# Molecular typing of breast cancer in Northern Henan Province

**DOI:** 10.1101/2021.07.28.21261239

**Authors:** Hui Zhao, Haibin Ma, Bangze Chen, Yahui Li, Junzheng Yang

## Abstract

**Objective:** Investigating and analyzing the clinical features of breast cancer patients in Northern Henan Province, measuring expression of biomarkers (ER, PR, HER2 and Ki-67) and classified the molecular typing of breast cancer patients, to understand the molecular typing distribution and correlation between biomarkers of breast cancer patients in North Henan Province, which may provide the information for the local oncologist to make sound treatment plans.

**Methods:** We collected the clinical data of breast cancer patients in Xinxiang Central Hospital from 2016 to 2021, those data was classified by gender and pathological types of breast cancer patients; and we also measured and analyzed the expression of breast cancer related biomarkers (ER, PR, HER2 and Ki-67) by immunohistochemistry, and based on expression of these biomarkers, the molecular typing of breast cancer were also classified.

**Results:** 3210 cases breast cancer patients were collected in this study; there were 3205 female patients and 5 male patients, accounting for 99.84% and 0.16% in total breast cancer patients, respectively. Classification according to pathological conditions of breast cancer patients, there were 2761 cases patients with invasive ductal carcinoma, accounting for 86.01% in total breast cancer patients, and then mucinous adenocarcinoma (109/3210, 3.40%), lobular carcinoma (106/3210, 3.30%), ductal carcinoma in situ (75/3210, 2.34%), papillary carcinoma (61/3210, 1.90%), intraductal carcinoma (40/3210, 1.25%), myeloid carcinoma (27/3210, 0.84%); There were also including some rare breast cancer types including cribriform carcinoma (6/3210, 0.19%), lymph node metastasis (7/3210, 0.22%), occult breast carcinoma (5/3210, 0.14%), invasive carcinoma (5/3210, 0.14%), squamous cell carcinoma (3/3210, 0.09%), fibroadenoma (3/3210, 0.09%), pleomorphic carcinoma (2/3210, 0.06%). Classification according to molecular typing of breast cancer, the number of breast cancer patients with Luminal A type [ER(+)/PR(+) HER2(-)Ki67<14%] were 207 cases, accounting for 6.45% in total breast cancer patients, the number of breast cancer patients with Luminal B type I [ER(+)/PR(+) HER2(-)] were 243 cases, accounting for 7.57% in total breast cancer patients, the number of breast cancer patients with Luminal B type II [(ER(+)/PR(+)HER2(+) any Ki67] was 254 cases, accounting for 7.91% in total breast cancer patients, and the number of Triple-negative breast cancer (TNBC) were 390 cases, accounting for 12.15% in total breast cancer patients.

The average expression rate of Ki-67 in ER (+) and/or PR (+) breast cancer patients was 20.39+27.33%, while the average expression rate of Ki-67 in ER(-)/PR(-) breast cancer patients was 36.35%+30.14%, and the difference between two patients was significant (p=0.0021); the average expression rate of Ki-67 in HER2 positive breast cancer patients was 23.01%+21.96%, the average expression rate of Ki-67 in HER2 negative breast cancer patients was 29.44%+24.16%, and there was no significant difference between the two groups (P=0.2589).

The main treatment methods of breast cancer patients in Northern Henan Province were antitumor drugs and chemotherapy, the results showed that 87.29% patients were treated by chemotherapy; and high frequency anti-tumor drugs used for breast cancer treatment were Epirubicin (1527/3210, 47.57%), Cyclophosphamide (1172/3210, 36.51%),Paclitaxel (1141/3210,35.55%), Tamoxifen (912/3210, 28.41%).

**Conclusions:** The main pathological type of breast cancer are invasive ductal carcinoma, and the main treatment methods of breast cancer patients in Northern Henan Province were antitumor drugs and chemotherapy. In the four kinds of molecular typing of breast cancer, the incidence rate of TNBC is highest compared with Luminal B type II[(ER(+)/PR(+)HER2(+) any Ki67], Luminal B type I [ER(+)/PR(+)HER2(-)] and Luminal A type [ER(+)/PR(+) HER2(-) Ki67<14%]; these results may provide some suggestions for the local oncologist.

## 1. Introduction

Breast cancer is one of the main causes for threatening women’s health worldwide. There is a high mortality rate no matter in developed countries or developing countries^[1]^. Compared with other countries, the incidence of breast cancer in China is relatively low; but with the environmental change and economic development, the number of breast cancer patients in China’s city and rural areas has been increasing in the past several years, diagnosis and treatment of breast cancer is becoming an issue that needs to attention.

Breast cancer is a highly heterogeneous cancer disease at the molecular level. With the development of science and technology and the in-depth study of breast cancer, scientists have found that some biomarkers such as estrogen receptor (ER), progesterone receptor (PR), human epidermal growth factor receptor-2 (HER2), and Ki-67 (proliferation marker) are not only highly associated with tumor growth, invasion, metastasis, recurrence, and they also has important guiding significance in the prediction and treatment of breast cancer^[2]^. Some articles have demonstrated that ER positive patients are generally treated with hormone therapy, and has good therapeutic effect; overexpression of HER2 in breast cancer patients is often closely related to invasive tumor types and poor clinical outcomes^[3]^; Ki-67, as a nuclear protein associated with cell proliferation, can be used as a prognostic marker for multiple malignancies including breast cancer^[4]^. Therefore, molecular typing of breast cancer is not only an important reference for personalized treatment of breast cancer, but also a good development direction for breast cancer treatment.

Breast cancer is usually divided into four subtypes according to breast cancer molecular typing method, including Luminal A type [ER(+)/PR(+)/HER2(-) Ki67<14%], Luminal B type I (ER(+)/PR(+)HER2(-)/Ki67>14%], Luminal B type II [ER(+)/PR(+)/HER2(+) any Ki67], and TNBC[(ER(-)/PR(-)/HER2(-)]. Luminal A type breast cancer, also known as hormone dependent breast cancer, is sensitive to endocrine therapy with good prognosis, but poor sensitivity to chemotherapy; is not sensitive to endocrine therapy, but endocrine therapy has a better survival rate for Luminal B type breast cancer. TNBC is the most studied subtype in breast cancer, it is not sensitive to endocrine therapy and HER2 treatment, and the five year survival rate is the lowest in the four kinds of subtypes of breast cancer.

In this study, we collected 3210 cases of breast cancer in Northern Henan Province, classified the data by gender and pathological types of breast cancer patients, and we also measured and analyzed the expression of breast cancer related biomarkers (ER, PR, HER2 and Ki-67) by immunohistochemistry, based on expression of these biomarkers, the molecular subtypes of breast cancer were also classified, which will lay the foundation for prevention and treatment of breast cancer in North Henan Province.

## 2 Research subjects

The clinical information of 3210 breast cancer patients were collected from 2016 to 2021 at Xinxiang Central Hospital in Northern Henan Province. The medical information including gender, expression of ER and PR, Ki-67 and HER2 were measured and recorded. The study was approved by Ethics Committee of Xinxiang Central Hospital. The informed consent was obtained from all patients. The procedures performed in this study involving human participants were in accordance with the Helsinki Declaration of 1975, as revised in 2008.

## 3 Research Methods

In this article, we collected the clinical data of 3210 breast cancer patients in Xinxiang Central Hospital from 2016 to 2021, the data was classified by gender and pathological types of breast cancer patients; and we also measured and analyzed the expression of breast cancer related biomarkers (ER, PR, HER2 and Ki-67) by immunohistochemistry, and based on expression of these biomarkers, the molecular subtypes of breast cancer were also classified.

Expression of ER, PR, HER-2 and Ki-67 was detected by immunohistochemistry staining, tissue species were formalin fixed, dehydrated in alcohol and embedded in paraffin. Immunostaining was performed according to the streptavidin-biotin peroxidase complex method, using monoclonal antibodies against ER, PR, HER-2 and Ki-67. Briefly, tissue species were cut into a thickness of 4-5μm, and dried 1 hour at 60°C in a fresh slide. And then All the slides were deparaffinized in xylene, rehydrated through alcohol, and washed in phosphate-buffered saline. This buffer was used for all subsequent washes. slide for ER, PR, HER-2 and Ki-67 detection were heated in a microwave oven twice for 5 min in citrate buffer (pH 6.0). Monoclonal antibodies (at a dilution of 1:500) ER, PR, HER-2 and Ki-67 were used respectively as the primary antibody and incubated overnight at room temperature followed by a conventional streptavidin peroxidase method. Signals were developed with 3, 30-diaminobenzidine for 5 minutes and counter-stained with hematoxylin. The results were scored for the positive signals in tissue cells. For ER and PR scoring, >1% positive staining cells in the breast cancer specimens were considered ER or PR positive expression. For HER-2 scoring, scores 0 and 1 were considered as negative expression, 2 and 3 were considered as positive expression. Ki-67 proliferation level was determined by the percentage of positive cells among the tested cells. For each slide, the pathological testing results were reviewed by two qualified pathologists for interpretation and diagnosis.

## 4 Statistical Analysis

Data are reported as the mean±SD. t-test or Fisher test was used to analyze the difference between different disease subtypes, P>0.05 was considered statistical difference.

## 5 Results

A total of clinical information of 3210 breast cancer patients were collected in Xinxiang central hospital from 2016 to 2021. There were 3205 female breast cancer patients and 5 male breast cancer patients, accounting for 99.84% and 0.16% in total breast cancer patients, respectively; Classification according to pathological conditions of breast cancer patients, there were 2761 cases patients with invasive ductal carcinoma, accounting for 86.01% in total breast cancer patients, and then mucinous adenocarcinoma (109/3210, 3.40%), lobular carcinoma (106/3210, 3.30%), ductal carcinoma in situ (75/3210, 2.34%), papillary carcinoma (61/3210, 1.90%), intraductal carcinoma (40/3210, 1.25%), myeloid carcinoma (27/3210, 0.84%); There were also including some rare pathological types of breast cancer including cribriform carcinoma (6/3210, 0.19%), lymph node metastasis (7/3210, 0.22%), occult breast carcinoma (5/3210, 0.14%), invasive carcinoma (5/3210, 0.14%), squamous cell carcinoma (3/3210, 0.09%), fibroadenoma (3/3210, 0.09%), pleomorphic carcinoma (2/3210, 0.06%) (Table 1).

**Table 1.** Pathological type distribution of breast cancer in 3210 cases of breast cancer patients in North Henan Province

| Breast cancer type | Cases | Proportion |
| --- | --- | --- |
| Invasive ductal carcinoma | 2761 | 86.01% |
| Mucinous adenocarcinoma | 109 | 3.40% |
| Lobular carcinoma | 106 | 3.30% |
| Ductal carcinoma in situ | 75 | 2.34% |
| Papillary carcinoma | 61 | 1.90% |
| Intraductal carcinoma | 40 | 1.25% |
| Medullary Breast Carcinoma | 27 | 0.84% |
| Cribriform carcinoma | 6 | 0.19% |
| Metastatic carcinoma of lymph node | 7 | 0.22% |
| Occult breast cancer | 5 | 0.16% |
| Invasive carcinoma | 5 | 0.16% |
| Squamous cell carcinoma | 3 | 0.09% |
| breast fibroadenoma | 3 | 0.09% |
| Pleomorphic carcinoma | 2 | 0.06% |
| Total | 3210 | 100.00% |

Subsequently, according to the results of immunohistochemistry, we classified and summarized biomarkers expression of ER, PR, HER2, and Ki-67 in 3210 cases breast cancer patients. The number of ER positive and ER negative breast cancer patients were 2379 cases and 831 cases, accounting for 74.11% and 25.89% in the total number of breast cancer patients, respectively; the number of PR positive and PR negative breast cancer patients were 2334 cases and 876 cases, accounting for 72.71% and 27.39% in the total number of patients respectively; the number of HER2 positive and breast cancer patients were 2561 cases and 649 cases, accounting for 79.78% and 20.22% in the total number of breast cancer patients respectively. The number of Luminal A type [ER(+)/PR(+)HER2(-)Ki67<14%] breast cancer patients were 207 cases, the number of Luminal B type I [ER(+)/PR(+)HER2(-)Ki67>14%] breast cancer patients was 243 cases, and the number of Luminal B type II [ER(+)/ PR(+)HER2(+) any Ki67] patients with breast cancer were 254 cases, the number of TNBC was 390 cases, accounting for 6.45%, 7.57%, 7.91%,12.15% in the total number of breast cancer patients, respectively (Table 2 and Table 3).

**Table 2.** Distribution of ER, PR and HER2 in 3210 breast cancer patients in North Henan Province

| in North Henan Province |  |  |  |
| --- | --- | --- | --- |
| Marker | Type | Cases | Proportion |
| ER | Positive | 2379 | 74.11% |
|  | Negative | 831 | 25.89% |
| PR | Positive | 2334 | 72.71% |
|  | Negative | 876 | 27.39% |
| HER2 | Positive | 2561 | 79.78% |
|  | Negative | 649 | 20.22% |

**Table 3.**
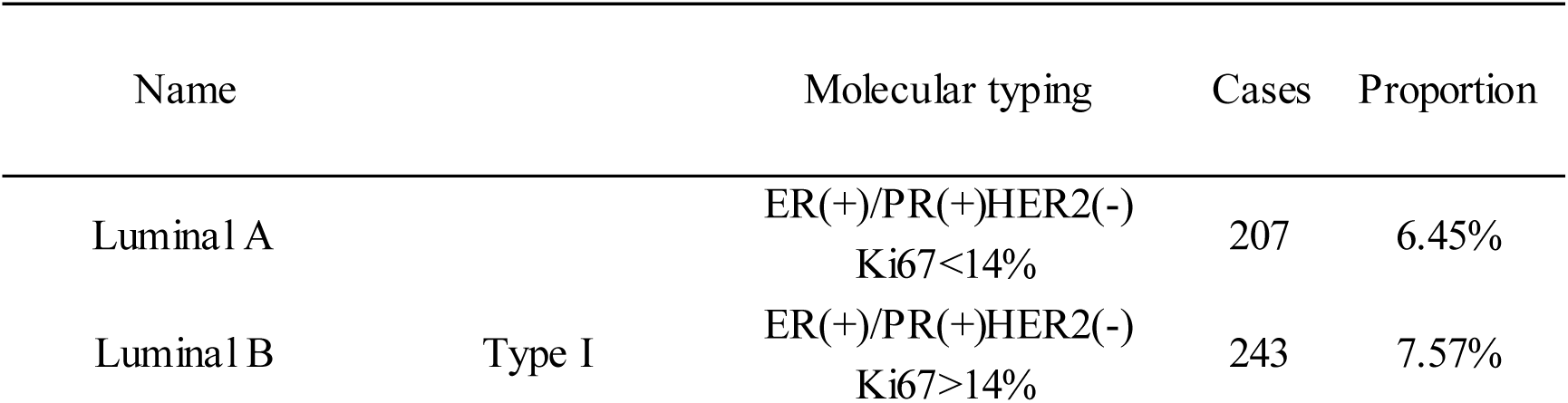
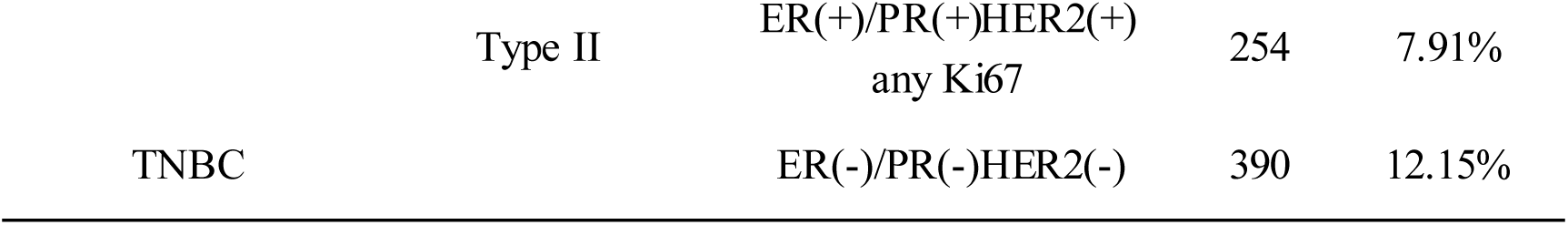
Molecular typing distribution in 3210 cases of breast cancer in North Henan Province

And then we studied the biomarkers relationship among ER, PR and Ki-67. The results showed that the number of ER(+) and/or PR(+) breast cancer patients was 2112 cases, accounting for 65.79% in the total number of breast cancer patients; the number of ER(-)/PR(-) breast cancer patients was 633 cases, accounting for 19.76% in the total number of breast cancer patients; the average expression rate of Ki-67 in ER(+) and/or PR(+) breast cancer patients was 20.39+27.33%, while the average expression rate of Ki-67 in ER(-)/PR(-) breast cancer patients was 36.35%+30.14%, there was the statistically significant between two groups (p=0.0021). We also analyzed relationship between HER2 and Ki-67. The number of HER2(+) breast cancer patients was 2561 cases, accounting for 79.78% in the total number of breast cancer patients; the number of HER2(-) breast cancer patients was 649 cases, accounting for 21.22% in the total number of breast cancer patients. The average expression rate of Ki-67 in HER2(+) breast cancer patients was 23.01%+21.96%, the average expression rate of Ki-67 in HER2(-) breast cancer patients was 29.44%+24.16%, and there was no significant difference between the two groups (P=0.2589) (Table 4).

**Table 4.** The correlation among biomarkers ER, PR and Ki-67 in breast cancer patients in Northern Henan Province

| Biomarkers | Cases | Ki-67 Level (%) (Mean±SD) | P-value |
| --- | --- | --- | --- |
| ER(+)and/or PR(+) | 2112 (65.79%) | 20.39±27.33% | 0.0021 |
| ER(-)/PR(-) | 633(19.76%) | 36.35% ±30.14% |  |
| HER2(+) | 2561(79.78%) | 23.01% ±21.96% | 0.2589 |
| HER2(-) | 649(21.22%) | 29.44% ±24.16% |  |

Finally, we summarized the treatment methods for breast cancer patients who visited the hospital in the Northern Henan Province, the results showed that 87.29% of breast cancer patients has been treated by chemotherapy; the highest frequency of antitumor drugs used was Epirubicin (1527/3210, 47.57%), Cyclophosphamide (1172/3210, 36.51%), Paclitaxel (1141/3210, 35.55%), Tamoxifen (912/3210). 28.41%) (Table 5).

**Table 5.** Treatment of breast cancer patients in North Henan Province

| Types | Cases | Proportion in 3210 patients |
| --- | --- | --- |
| Chemotherapy | 2802 | 87.29% |
| Cyclophosphamide | 1172 | 36.51% |
| Epirubicin | 1527 | 47.57% |
| Cisplatin | 153 | 4.77% |
| Carboplatin | 252 | 7.85% |
| Paclitaxel | 1141 | 35.55% |
| Docetaxel | 714 | 22.24% |
| Vinorelbine | 160 | 4.98% |
| Gemcitabine | 73 | 2.27% |
| herceptin | 250 | 7.79% |
| Fluorouracile | 248 | 7.73% |
| Methotrexate | 62 | 1.93% |
| Tamoxifen | 912 | 28.41% |
| xeloda | 228 | 7.10% |
| Etoposide | 19 | 0.60% |
| AUSTIN | 7 | 0.22% |
| Pemetrexed | 9 | 0.28% |
| Exemestane | 253 | 7.89% |
| Torremifen | 310 | 9.66% |
| Letrozole | 271 | 8.44% |
| Anastrozole | 508 | 15.83% |
| Norred | 130 | 4.05% |
| Fulvestrant | 6 | 0.19% |
| Methylprogesterone | 24 | 0.75% |
| Bisphosphate | 97 | 3.02% |
| Lapatinib | 8 | 0.25% |

## 6 Discussion

We collected clinical data of 3210 breast cancer patients in Xinxiang Central Hospital from 2016 to 2021. There were 3205 female patients and 5 male patients, accounting for 99.84% and 0.16% respectively; It is found that the existence of male breast cancer in the Northern Henan Province, due to the structural and physiological particularity of male breast, medical staff should pay attention to male psychological factors while treatment. Then we classified breast cancer patients according to the pathological conditions of patients, found that 2761 cases patients were invasive ductal carcinoma, accounting for 86.01% in the total number of breast cancer patients, 109 cases patients were mucinous adenocarcinoma, 106 cases patients were lobular carcinoma, 75 cases patients were ductal carcinoma in situ; it also existed some rare types of breast cancer: cribriform carcinoma (6/7210, 0.19%), lymph node metastasis (7/3210, 0.22%), occult breast cancer (5/3210, 0.16%), invasive carcinoma (5/3210, 0.16%), squamous cell carcinoma (3/3210, 0.09%), fibroadenoma (3/3210, 0.09%), pleomorphic carcinoma (2/3210, 0.06%). Those data demonstrated that invasive ductal carcinoma was mainly pathological type of breast cancer in Northern Henan Province, and multiple types of breast cancer subtypes coexisted, which suggest that clinical staff should judge the disease from multiple perspectives and factors in diagnosis and treatment to improve the treatment effect.

It was reported that the response rate of endocrine therapy in ER(+)/HER2(+) breast cancer patients can be reduced from 84% to 20%, the response rate of endocrine therapy in ER(-)/HER2(+) breast cancer patients can be reduced from 27% to 0; and expression of HER2 could inhibit the expression of hormone receptor. Therefore, the treatment of Luminal B type breast cancer usually requires endocrine therapy plus chemotherapy^[5, 6]^. Because breast cancer with HER2 overexpression is more sensitive to Taxol chemotherapy, chemotherapy combined with biological targeting drugs has a good therapeutic effect on breast cancer with HER2 overexpression^[7, 8]^. In this article, we measured and analyzed the expression of ER, PR, HER2 and Ki-67 by immunohistochemistry. We found that the number of ER positive breast cancer patients PR positive breast cancer patients and HER2 positive breast cancer patients, accounting for 74.11%, 72.71% and 79.78% in the total breast cancer patients respectively. Luminal A type [ER(+)/PR(+)HER2(-)Ki67<14%] of breast cancer patients, Luminal B type I [ER(+)/PR(+)HER2(-)Ki67>14%] breast cancer patients Luminal B type II [ER(+)/PR(+)HER2(+) any Ki67] breast cancer patients, and TNBC were accounting for 6.45%, 7.57%, 7.91%,12.15% in the total number of breast cancer patients, respectively. Because the four kinds of molecular typing of breast cancer are different in their tolerance and selectivity to the current treatment methods, it is usually necessary to conduct a relative treatment based on molecular typing, suggesting that the clinical staff should proceed with the diagnosis and treatment according to the molecular typing of breast cancer and the actual situation of the patients.

At the same time, we studied the relationship among ER, PR, HER2 and Ki-67. The average expression rate of Ki-67 in ER positive and/or PR positive breast cancer patients was 20.39+27.33%, while the average expression rate of Ki-67 in patients with ER(-)/PR(-) breast cancer was 36.35%+30.14%, there was a significant difference between the two groups (p=0.0021), the data showed that there was a correlation among ER, PR and Ki-67. The average expression rate of Ki-67 in HER2 positive breast cancer patients was 23.01%+21.96%, and the average expression rate of Ki-67 in HER2 negative breast cancer patients was 29.44%+24.16%, there was no significant difference between the two groups, therefore there was no correlation between HER2 and Ki-67 (P=0.2589). Those results suggests that clinicians should consider the expression changes of biomarkers such as ER, PR, HER2 and Ki-67 in the diagnosis and treatment of breast cancer.

Finally, we summarized the treatment methods of breast cancer patients in the Northern Henan Province, and found that 87.29% of breast cancer patients were treated by chemotherapy. The highest frequency of antitumor drugs used for breast cancer treatment was Epirubicin (1527/3210, 47.57%), Cyclophosphamide (1172/3210, 36.51%), Paclitaxel (1141/3210, 35.55%), Tamoxifen (912/3210, 28.41%). indicated that treatment of breast cancer in the northern Henan Province were mainly chemotherapy and antitumor drugs.

## 7 Conclusion

In this article, we summarized the clinical features of 3210 cases breast cancer patients in North Henan Province from 2016 to 2021. It is found that invasive ductal carcinoma is the main pathological type of breast cancer in North Henan Province. In the four kinds of molecular typing of breast cancer, TNBC had a highest incidence rate compared with the other types of breast cancer. it is recommended that reasonable diagnosis and treatment measures should be taken to prevent breast cancer and to improve the therapeutic effect.

## Data Availability

The data used to support the findings of this study are available from the corresponding author upon request.

## 9 Acknowledgements

None.

